# Morphometric and molecular analysis of Schistosomes eggs recovered from human urines in communities along the shore-line of Oyan-dam in Ogun State, Nigeria

**DOI:** 10.1101/2022.04.11.22273687

**Authors:** Adedotun Ayodeji Bayegun, Olaitan Olamide Omitola, Uche Cynthia Umunnakwe, Adedayo Foluke Akande, Olaoluwa P. Akinwale, Hammed Oladeji Mogaji, Kehinde O. Ademolu, Pam Vincent Gyang, Nnayere Simon Odoemene, Uwem Friday Ekpo

## Abstract

**Background:** There are growing concerns that communities characterized with surface water, where both humans and livestock interact for agricultural, domestic, cultural, and recreational purposes, are likely to support hybridization between schistosome species infecting humans and livestock. This study therefore investigated the possible human infections with hybrid schistosomes in four schistosomiasis endemic communities along the banks of Oyan dam in Ogun State, Nigeria.

**Methods:** Human urine samples were collected in Imala-Odo, Abule-Titun, Apojula and Ibaro-Oyan communities. Recovered eggs were counted, photographed, and measured with IC Measure™ for Total Length, Maximum Width, and a ratio of egg shape. Eighty-seven unusual *Schistosoma* eggs shaped were molecularly characterised by PCR amplification of *Schistosoma* specific *Dra1* gene. The amplicons were further subjected to PCR amplification of schistosome ITS-2 rDNA and right representative samples with varying gel band sizes were sequenced.

**Results:** A total of 1,984 *Schistosoma* eggs were analysed. The egg morphometrics were within the range of 70.90 - 262.30 μm and 30.10 - 102.60 μm for total length and width respectively. The length to width ratio was 1.60 - 4.06μm. Majority of the eggs have the typical round-to-oval shaped eggs (1345, 67.8 %), followed by eggs with atypical spindle-shaped (639, 32.2 %) and eggs without spines (22, 1.1 %). Egg morphotypes were significantly different (p = 0.017). PCR amplification of *Dra1* gene and ITS2 confirmed 54 (62.1%) of the eggs and 33 (61.1%) of *Schistosoma* origin. Sequencing of two of the DNA samples produced sequences similar to vertebrate *S. magrebowiei* (accession number UZAI01000474.1) and Asian *S. japonicum* (accession number SKCS01001458.1).

**Conclusion:** These findings suggest possibly that hybrids schistosome may be circulating in the human population in the study areas.

**Author summary:** Human schistosomiasis is one of the most common neglected tropical diseases in Africa. The disease is caused by a water-borne trematode parasite, and transmission among human requires contact with infested water bodies. There are growing concerns that communities characterized with infested surface water, where both humans and livestock interact are likely to support hybridization between schistosome species infecting humans and livestock. We therefore screened human urine samples using morphological and molecular methods, for the presence of hybrid schistosomes in four communities along the banks of Oyan dam in Ogun State, Nigeria. Our findings show the possible occurrence of hybrid schistosomes in the study area, which call for urgent public health interventions.

## Introduction

Human schistosomiasis is one of the most prominent neglected tropical diseases (NTDs) in Africa [1]. The disease is caused by water-borne trematode parasites of the genus Schistosoma, and is widely distributed in 78 countries with about 206 million cases, 24,000 deaths and 2.5 million disability adjusted life years [2]. There are six species of the genus Schistosoma infecting humans worldwide, with two major species in Africa; *S. haematobium* and *S. mansoni*. The former is responsible for most of the morbidity in Africa, with the adult parasite inhabiting the vesicular and pelvic venous plexus of the bladder and causing urogenital schistosomiasis [3]. The pathologies associated with this species is dependent on the severity of infection, migration of the worms and inflammatory responses to the presence of the eggs [3,4].

Largely, schistosomiasis is a focal disease [5], which thrives in rural and marginalized urban populations that share proximities with surface waterbodies, and are characterized by inadequate or poor access to water, sanitation and hygiene (WASH) facilities [6]. In such areas, contact activities with surface waterbodies ranges from bathing, washing of clothes, swimming and playing are not uncommon [6]. These activities support the transmission cycle of the parasites, with infestation of waterbodies through open urination or defecation by an infected resident, and subsequent infection of other residents through other domestic contact purposes. Control and elimination for schistosomiasis have therefore focused on mass administration (MDA) of praziquantel to children between ages 5 and 15 years in endemic communities [2], with possible complementary provision of WASH interventions to promote behavioural change [7]. However, in endemic communities, with infested surface waterbodies, where both humans and livestock interact, there are growing concerns on the hybridization of closely related species of humans (*S. haematobium*) and cattle (*S. bovis*) in the laboratory and the wild [8]. The emergence of hybrid species has raised significant concerns for schistosomiasis control effort [9].

Nigeria has the highest burden of schistosomiasis in Africa [1], with the disease been endemic across all 36 States in the country [10]. Around certain transmission foci, precisely, communities situated along the banks of Oyan-dam in the southwestern part of the country, prevalence can reach as high as 90% [11]. The communities (Abule-Titun, Apojola, Ibaro and Imala-Odo) have remained highly endemic since 1991 despite ongoing interventions [12]. Predominant water contact activities like fishing, farming, bathing, swimming, drinking, washing clothes or kitchen utensils and fetching of water from infested surface waterbodies are common [11,12]. Furthermore, livestock farming is one of the most common occupation of the populace, and this allows interaction between cattle with humans along the banks of the dam that surrounds these communities (Fig 1). Since, *S. bovis* has been previously reported as the predominant livestock species in the country [13], we therefore hypothesize that *Schistosoma* hybridization and zoonotic transmission may be ongoing in Nigeria in obscurity. This baseline study was therefore conducted to characterize *Schistosoma* eggs obtained from in human urine to screen for possible zoonotic and hybrid schistosomes using morphometric and molecular methods.

**Fig 1.**
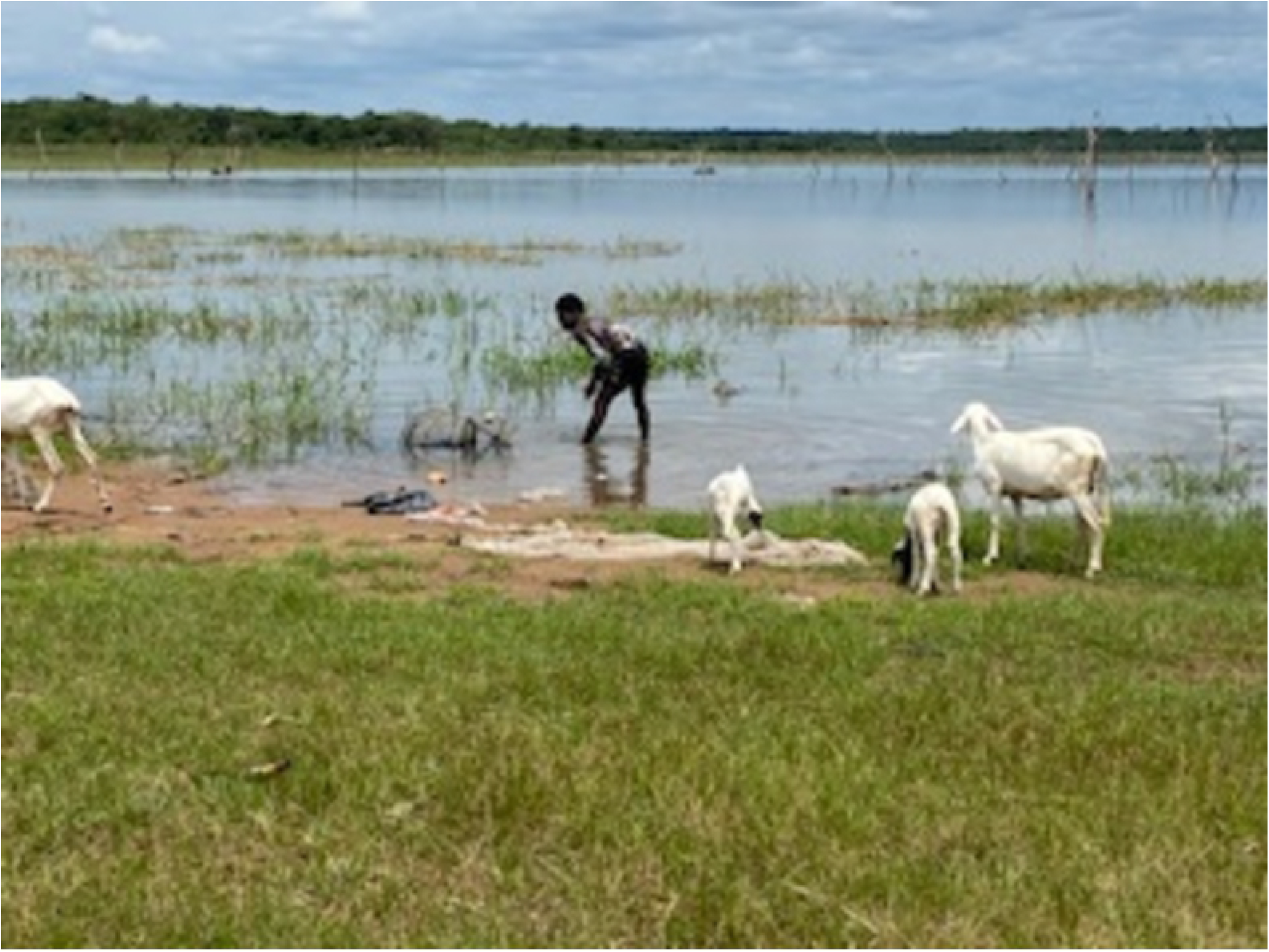
Sharing of common water source by humans and cattle at Apojola community.

## Methods

### Ethical Statements and Consideration

Ethical clearance for this study (SHA/RES/VOL.4/154) was obtained from Ogun State Hospital, Ijaiye health ethics review board. Pre-survey contact/advocacy meeting was made to study communities to obtain consents from the community leaders after explaining the objectives of the research to them. Communities willing to participate in the study completed written consent forms. Subsequently, parents and guardians/caregivers were also informed about the study through community meetings organized by the consenting representatives. Parents were asked to provide parental consents to their children by completing another consent form, in addition to an assent form which was completed by children below 16 years of age. Children whose parent did not agree to the study procedures were not recruited in the research.

### Study communities

This study was conducted in four communities along the shoreline of Oyan River Dam in Ogun State, Nigeria. Ogun State is one of the 36 States in the Federal Republic of Nigeria, located in the southwestern region of the country (Fig 2). It covers a total area of 16,409.26 sq. km between latitude 6.2°N and 7.8°N and longitude 3.0°E and 5.0°E. The State has 20 Local Government Areas (LGA) comprising 236 political wards. The greater proportion of the State lies in the tropical rainforest zone, while the far northern part has features of the Guinea Savannah. In the early 90s, the Oyan River Dam was established, with its shorelines around four major communities; Imala-Odo and Ibaro Oyan in Abeokuta North LGA, and Apojola and Abule-titun in Odeda LGA (Fig 2). These communities have remained endemic for schistosomiasis since 1991 [12], with predominant occupations been fishing and livestock farming [11]. This in addition to poor access to improved WASH facilities have also promoted water contact activities such as bathing, swimming, drinking, washing clothes or kitchen utensils and fetching of water from surface waterbodies, and more importantly shared interaction between cattle with humans along the shoreline of the dam that surrounds these communities (Fig 1).

**Fig 2:**
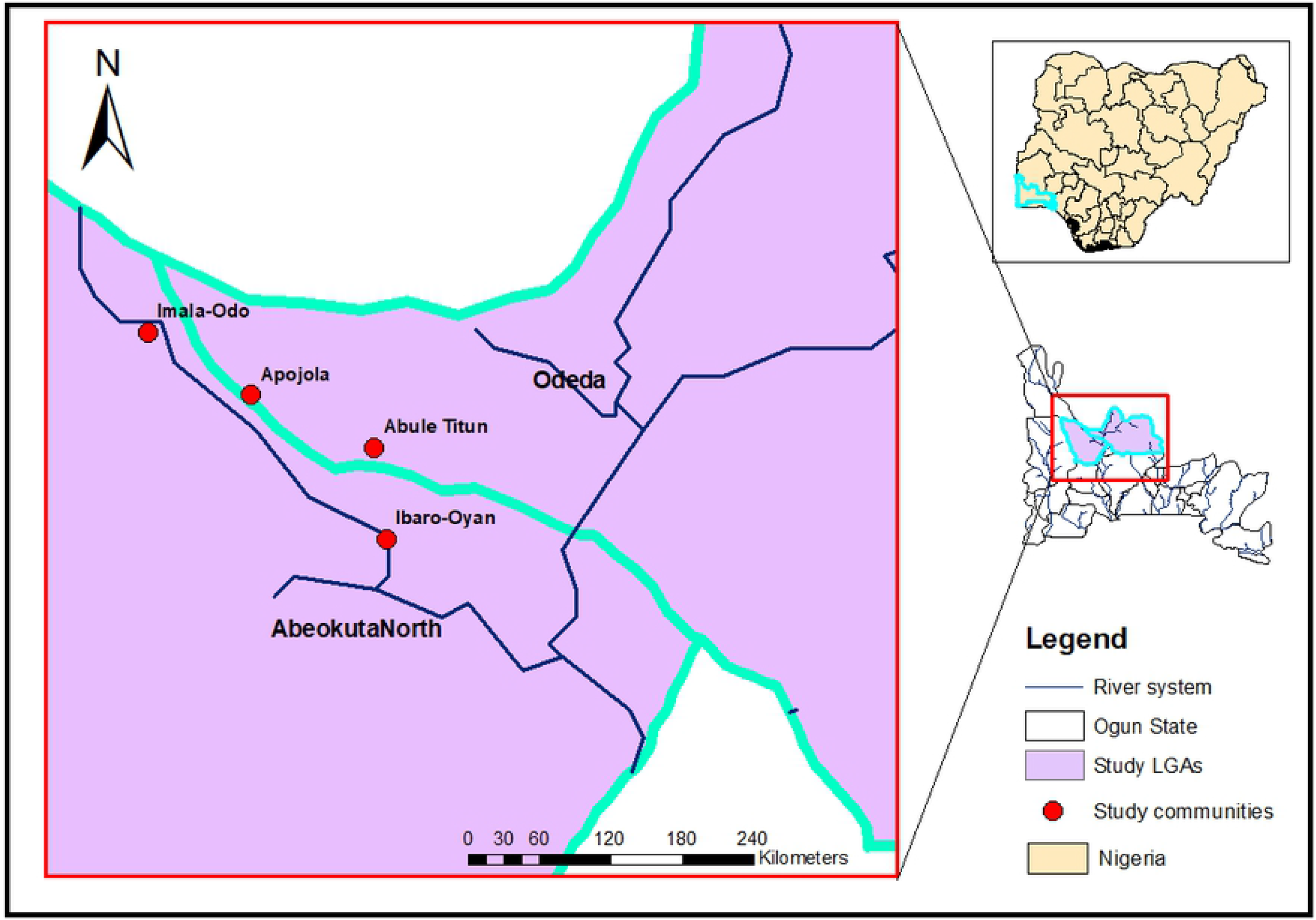
Map of Ogun State showing the study communities and river system.

### Sample size determination and recruitment of study participants

The sample size for this study was determined using the formula described by [14], 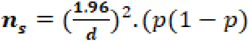 where n_s_ is the sample size, p is the existing prevalence in the study area, and d is the degree of accuracy. In determining the sample size, a prevalence of 47% [12], and a degree of accuracy of 5% was considered at 95% level of confidence. The minimum sample size determined therefore was 383 i.e., an average of 96 persons per community. The community were compact, and invitation to participate were sent to all residents through a local mobiliser. Recruitment and collection of samples was done at a central location and only residents who consented to the study procedures were enrolled into the study.

### Sample collection and examination

Urine samples were collected from consenting residents comprising of infants and preschoolers (1-4years), school-aged children (5-15 years) and those above 16-64 years across study communities between October and November, 2019. Age of participants were validated using birth-cards to avoid information bias. Three-hundred and eighty-four samples were collected in dark, sterile 30ml universal containers and preserved with 70% ethanol. Collections were made between 10.00 and 14.00 hours as recommended [15] and transported to the laboratory in iceboxes. Urine samples were processed using sedimentation techniques, and examined under the microscope for the presence of *S. haematobium* eggs. A total of 219 (57.0%) of the samples examined were positive for *S. haematobium* eggs, and seperated for subsequent screening using morphometric and molecular methods.

### Morphometric analysis of *S haematobium* eggs

A total of 1984 eggs were examined using morphometrics methods. Microphotographs of the *Schistosoma* eggs and ova were taken using an AmScope MD130 1.3MP Digital Microscope (United Scope LLC., CA, USA) and the IC Measure™ (The Imaging Source Europe GmbH, Bremen, Germany) computer software was used to measure the total length (including the terminal spine) and the maximum width. The egg length/width ratio was subsequently computed. Qualitative characteristics such as unusual morphology was noted, and the presence or absence of the terminal spine was also recorded. The eggs were classified as typical if they have a round-to-oval shape or atypical if they have spindle/elongated shape [16,17]. Putative hybrid schistosomes were identified by their atypical egg-shape [16,18]. A total of 639 (32.2%) eggs were characterized with atypical shape.

### DNA extraction of the *Schistosoma* eggs

Eighty-seven (14%) of the atypical *Schistosoma* eggs were randomly selected for DNA extraction. The eggs were collected individually with a micropipette using a microscope and transferred to a 1.5ml reaction tube. To extract DNA, 150 μ l of the extraction buffer ([M Sodium chloride, 0.05M Tris-HCl, pH7.5, 1mM EDTA, 10% SDS], Proteinase K to a final concentration of 60 μ l/ml (0.6mg/extraction buffer of 100 ml) was added to each of the tubes, mix well using the pulse vortex machine and incubated for 2 hours at 52°C. After incubation, 150 μ l volume of phenol (pH 8.0) was added to the mixture and mixed gently for 10 minutes. The mixture was centrifuged for 10 minutes at 13000 rpm, the aqueous layer of the mixture was carefully removed into another 1.5ml reaction tube and 75 μ l volume of phenol and chloroform-isoamyl alcohol was added respectively. The mixture was pulse vortexed and centrifuged for 10 minutes respectively and the aqueous layer was removed into another 1.5ml reaction tube where 150 μ l of chloroform-isoamyl alcohol was added. Again, the mixture was pulse vortexed and centrifuged for 10 minutes respectively and the aqueous layer was removed into another 1.5ml reaction tube where 15 μ l volume of 3M sodium acetate (pH 5.2) and 375 μ l volume of ice-cold ethanol was added. The extracted DNA was precipitated overnight at -20°C. The DNA extraction procedure continued with centrifugation of the precipitated samples for 10 minutes at 13000 rpm. The pellets were washed twice with 200 μ l of 70 % ice-cold ethanol and for each wash, and centrifuged for 10 minutes at 13000 rpm. The pellets were dried and re-suspend in TE buffer (20 μ l) and stored at -20°C until used.

### PCR amplification of schistosome *Dra1* and ITS-2 subunit genes

The extracted DNA samples were subjected to PCR amplification of the schistosome *Dra1* repeat using forward primers 5’GATCTCACCTATCAGACGAAAC3’ and reverse primers, 5’TCACAACGATACGACCAAC 3’ [19]. The schistosome ITS-2 subunit (including most of the 5.8S gene and 40 bases of the 5’ of the 28S gene) was also amplified using two ‘‘universal primers’’ (STAB VIDA, Lda, FCT/UNL Ed. Departmental (Química/Ambiente), Lab 007) ITS-3 (5’-GCA TCG ATG AAG AACGCA GC-3’) and ITS-4 (5’
s-TCC TCC GCT TAT TGA TAT GC-3’) as described previously [20]. A 20μl PCR consisting of 4μl of the 5x FirePol Master Mix Ready to Load, and 5ng/μl of template DNA was performed for each. In the *Dra1* reactions, 0.25μl of the forward primer, 0.25μl of the reverse primer and 10.5μl of ddH_2_O were used while 0.1μM of ITS-3 and 0.1μM of ITS-4 primers,10.8 μl of ddH_2_O were used for the ITS2 reactions. *S. haematobium* DNA obtained from BEI Resources of the NIAID (NIH, USA) and distilled water were used as a positive and negative control, respectively. The amplification cycling conditions were initiation at 94°C for 4 minutes, denaturation at 94°C for 30 cycles for 1 minute, annealing at 50°C for 30 seconds, extension at 72°C for 45 seconds, final extension at 72 °C for 7 minutes and then held at 4°C. The PCR amplifications were carried out with the BioRad iCycler. Amplified products were visualized on 1.5% agarose gel with ethidium bromide stain and photographed with Gel Documentation and Analysis System (Clinx Science Instruments). Tandemly repeated bands were identified as positive *Dra1* amplicons while an ITS-2 DNA fragment of about 501 bp was considered positive. Amplicons with different band sizes for the ITS-2 rDNA were also selected for sequencing.

### Data analysis

Data collected were analysed using SPSS version 23.0 for Windows. Descriptive statistics and differences in proportions were tested using the Chi-square statistics, either for trend or independence, as appropriate. The number of eggs counted was transformed using lograthmic function (log (n+1)), to normalize the distribution of the residual values for statistical analyses. Differences between means across the study communities were tested using One-way analysis of variance (ANOVA). Morphometric data were exported from the IC Measure™ into Microsoft Excel for analysis. Statistical difference was set at 95 % confidence interval (p-value < 0.05).

## Results

### Demographic characteristics of study participants and infection status

A total of 384 participants were recruited, 198 (51.6%) males and 186 (48.4%) females, between the age group 1-4 years (112; 29.2%), 5-15 years (190; 49.5%) and 16-64 years (82; 21.4%). An overall prevalence of 219 (57.0%) was recorded, with majority of the infection among the participants from Ibaro-Oyan with a prevalence of 62.4%. Also, the prevalence of infection was higher in the males (31.8%) and in the 5-15 years age group (26.0%) (Table 1).

**Table 1:** Demographic characteristics and infection status of study participants.

|  | Communities |  |  |  |  |  |  |  |  |  |
| --- | --- | --- | --- | --- | --- | --- | --- | --- | --- | --- |
|  | Imala-Odo |  | Abule Titun |  | Apojola |  | Ibaro-Oyan |  | Total |  |
|  | NE (%) | NI (%) | NE (%) | NI (%) | NE (%) | NI (%) | NE (%) | NI (%) | NE (%) | NI (%) |
| **Sex** |  |  |  |  |  |  |  |  |  |  |
| Male | 55 (58.5) | 34 (36.2) | 51 (58.6) | 29 (33.3) | 61 (51.7) | 39 (33.1) | 31 (36.5) | 20 (23.5) | 198 (51.6) | 122 (31.8) |
| Female | 39 (41.5) | 21 (22.3) | 36 (41.4) | 18 (20.7) | 57 (48.3) | 25 (21.2) | 54 (63.5) | 33 (38.8) | 186 (48.4) | 97 (25.3) |
| Total | 94 (100.0) | 55 (58.5) | 87 (100.00) | 47 (54.0) | 118 (100.0) | 64 (54.2) | 85 (100.0) | 53 (62.4) | 384 (100.0) | 219 (57.0) |
| p-value |  | 0.525 |  | 0.663 |  | 0.042 |  | 0.819 |  | 0.064 |
| **Age group (in years)** |  |  |  |  |  |  |  |  |  |  |
| 1-4 | 29 (30.9) | 17 (18.1) | 22 (25.3) | 11 (12.6) | 32 (27.1) | 20 (16.9) | 29 (34.1) | 18 (21.2) | 112 (29.2) | 66 (17.2) |
| 5-15 | 53 (56.4) | 29 (30.9) | 49 (56.3) | 27 (31.0) | 50 (42.4) | 20 (16.9) | 38 (44.7) | 24 (28.2) | 190 (49.5) | 100 (26.0) |
| 16 -64 | 12 (12.8) | 9 (9.6) | 16 (18.4) | 9 (10.3) | 36 (30.5) | 24 (20.3) | 18 (21.2) | 11 (12.9) | 82 (21.4) | 53 (13.8) |
| Total | 94 (100.0) | 55 (58.5) | 87 (100.0) | 47 (54.0) | 118 (100.0) | 64 (54.2) | 85 (100.0) | 53 (62.4) | 384 (100.0) | 219 (57.0) |
| p-value |  | 0.436 |  | 0.906 |  | 0.027 |  | 0.988 |  | 0.165 |

**NE: Number Examined; NI: Number Infected**

### Morphotypes of *Schistosoma* eggs across the study areas

A total of 1,984 schistosome eggs from 219 (57.0%) infected individuals were photographed and measured. This comprised 605 (30.5%), 313 (15.8%), 775 (39.1%) and 291 (14.7%) eggs from Imala-Odo, Abule-titun, Apojola and Ibaro-oyan, respectively (Table 2). Three egg morphotypes were recorded in this study (Fig 3, 4 and 5). The majority of the schistosome eggs were of the typical round-to-oval shape (1345, 67.8%), and atypical elongated or spindle-shape (639, 32.2%). By location, 417 (68.9%), 192 (61.3%), 523 (67.5%) and 213 (73.2%) of the eggs recovered from Imala-Odo, Abule-titun, Apojola and Ibaro-oyan were round-to-oval shape, while 188 (31.1%), 121 (38.7%), 252 (32.5%) and 78 (26.8 %) were spindle-shaped respectively (Table 2). There was a significant variation in egg morphotypes across the study area (p = 0.017). Furthermore, spineless *Schistosoma* eggs (22, 1.1%), also co-existed with eggs recovered from Imala-odo, (8, 1.3 %), Abule-titun (5, 1.6 %), Apojola (5, 0.6 %) and Ibaro-oyan (4, 1.4 %).

**Table 2:** The morphotypes of Schistosoma eggs across the study areas.

| Communities | NEE | Egg Morphotypes |  |  |
| --- | --- | --- | --- | --- |
|  |  | Round-to-oval shape | Spindle shape | Spineless* |
| Imala-Odo | 605 (30.5) | 417 (68.9) | 188 (31.1) | 8 (1.3) |
| Abule-Titun | 313 (15.8) | 192 (61.3) | 121 (38.7) | 5 (1.6) |
| Apojola | 775 (39.1) | 523 (67.5) | 252 (32.5) | 5 (0.6) |
| Ibaro-oyan | 291 (14.7) | 213 (73.2) | 78 (26.8) | 4 (1.4) |
| Total | 1984 | 1345 (67.8) | 639 (32.2) | 22 (1.1) |
| p-value |  | 0.017 |  | 0.451 |

NEE; Number of eggs examined; \*Co-existing spineless *Schistosoma* eggs

**Fig 3.**
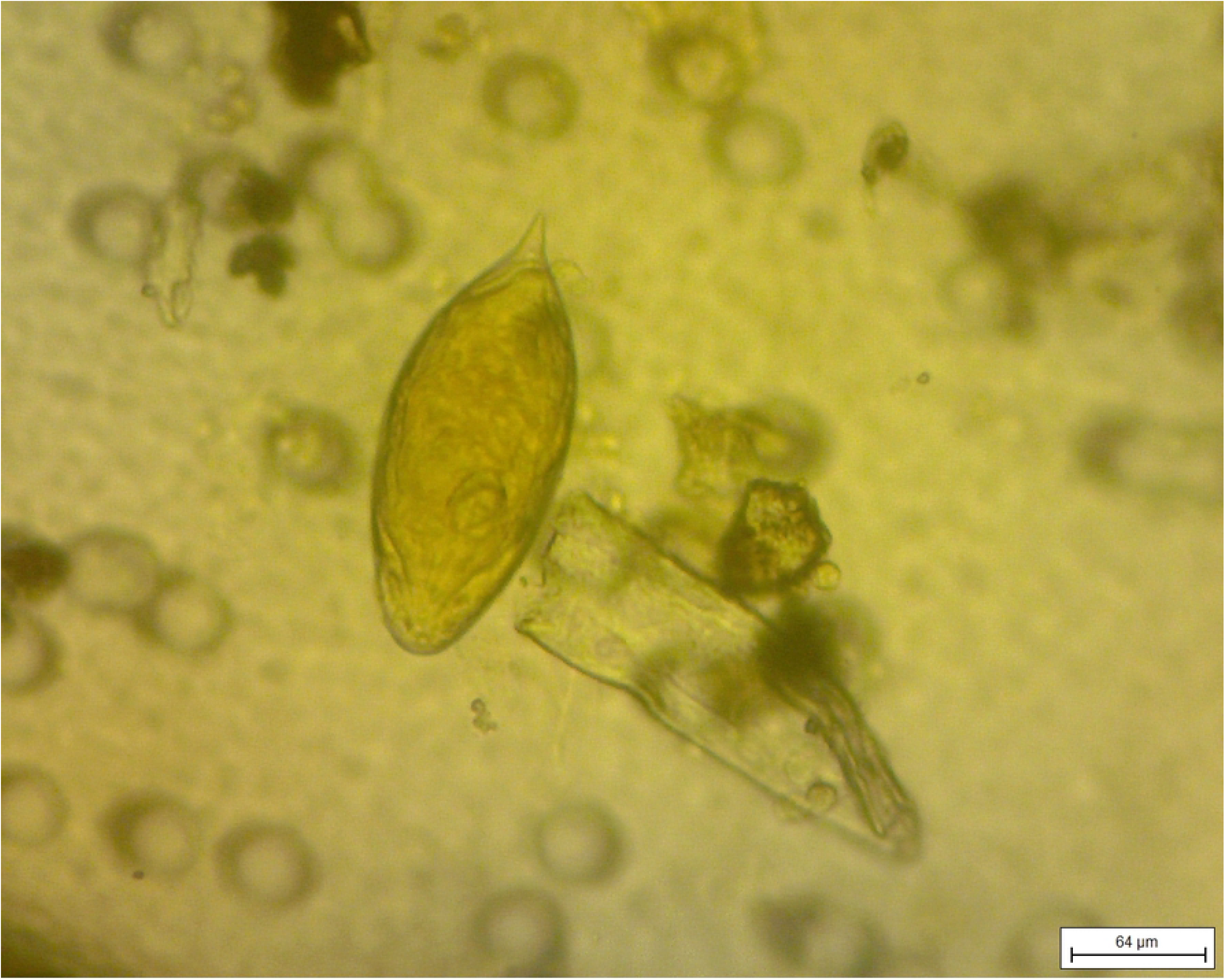
A typical Schistosoma egg with the terminal spine.

**Fig. 4.**
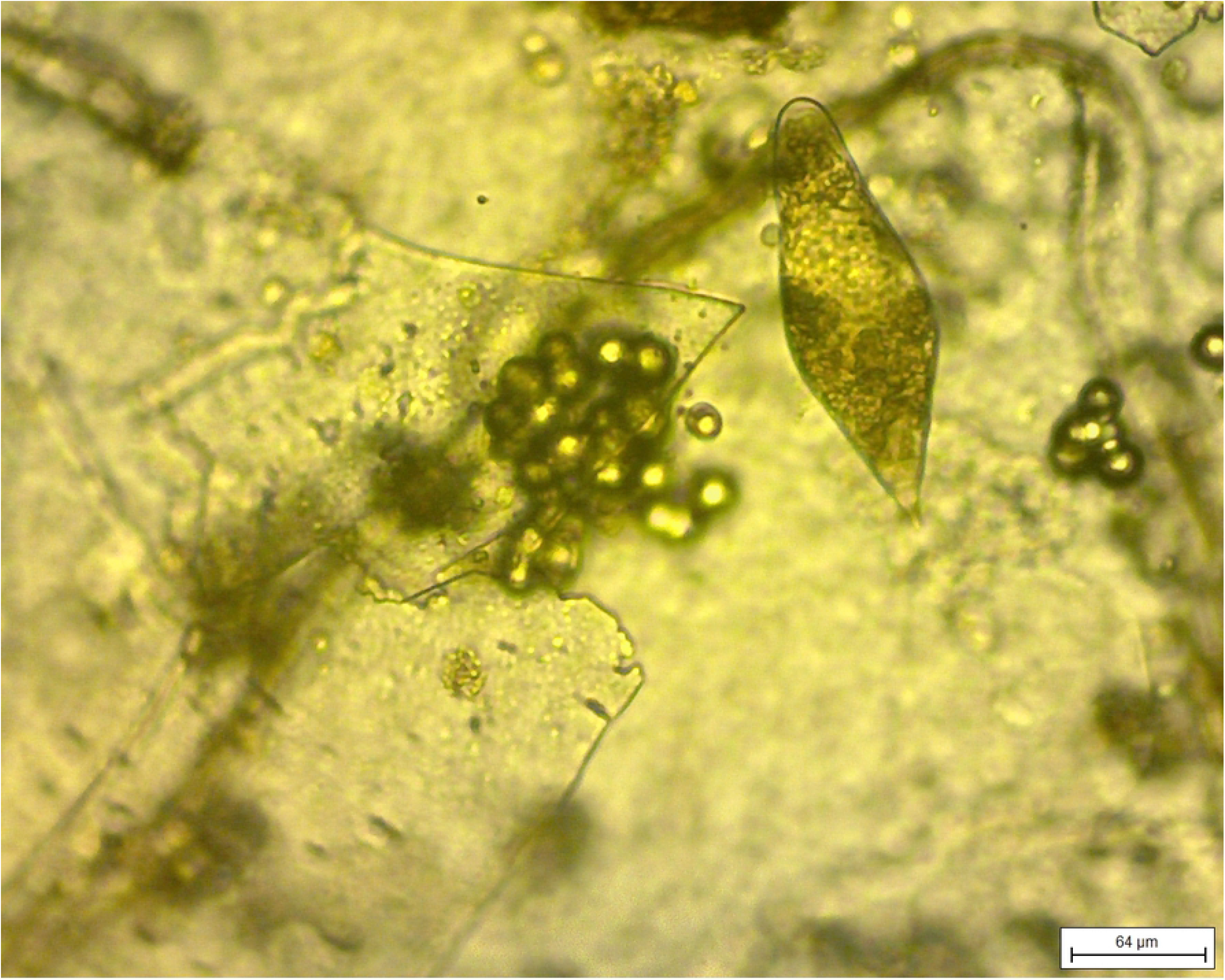
An Atypical spindle-shaped Schistosoma egg.

**Fig. 5.**
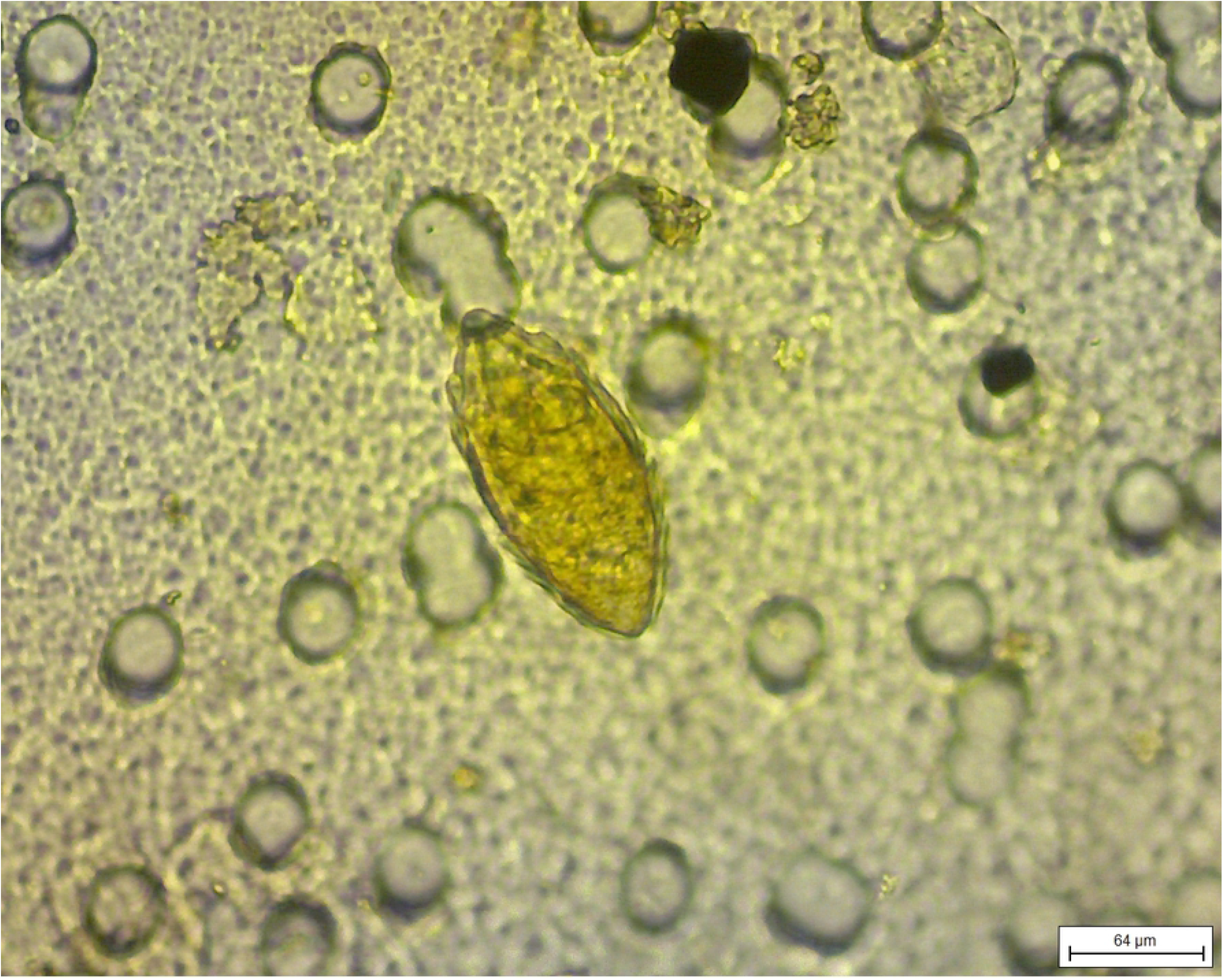
A spineless Schistosoma egg.

### Morphometrics of *Schistosoma* eggs across the study areas

Of the 1,984 *Schistosoma* eggs recovered from the human urine samples across the study areas, the total length ranged from 70.90 μm - 262.30 μm with a mean length of 176.97 μm (Table 3). The maximum width of the eggs was 102.60 μm with the minimum egg width of 30.10 μm and the mean egg width of 68.25 μm. Table 3 showed that the length/width ratio of the eggs had a range of 1.60 μm - 4.06 μm and the mean egg length/width ratio of 2.61 μm. Figure 6 shows the scatterplot distribution of the egg length against the egg width which represents an uneven distribution between the egg morphotypes from the areas. Only one positive schistosome egg was recovered from the sampled cattle, and the egg length, width and length/width was 330.10 μm, 139.50 μm and 2.40 respectively (Table 3).

**Table 3:** The morphometrics of Schistosoma eggs across the study areas (Range of Typical S. haematobium egg: 83-187 μm [19])

| Communities |  |  |  |  |  |  |  |  |  |  |  |
| --- | --- | --- | --- | --- | --- | --- | --- | --- | --- | --- | --- |
|  | Imala-Odo |  | Abule Titun |  | Apojola |  | Ibaro-Oyan |  | Total |  | p-value |
|  | X ± S.D | Min-Max | X ± S.D | Min-Max | X ± S.D | Min-Max | X ± S.D | Min-Max | X ± S.D | Min-Max |  |
| Length (µm) | 175.16±25.50 | 73.00-262.30 | 183.50±18.62 | 116.90-247.40 | 175.70±25.48 | 70.90-234.80 | 177.11±17.40 | 125.80-218.90 | 176.97±23.63 | 70.90-262.30 | 0.000 |
| Width (µm) | 67.12±9.39 | 33.40-95.50 | 71.15±8.08 | 46.20-95.50 | 67.09±9.51 | 30.10-97.50 | 70.55±6.95 | 48.90-102.60 | 68.25±9.08 | 30.10-102.60 | 0.000 |
| Length/width ratio (µm) | 2.63±0.31 | 1.60-3.92 | 2.60±0.32 | 1.65-3.81 | 2.64±0.32 | 1.72-4.06 | 2.52±0.28 | 1.65-3.33 | 2.61±0.31 | 1.60-4.06 | 0.000 |

285 **X: Mean value; S.D: Standard deviation; Min: Minimum value; Max: Maximum value**

**Figure 6:**
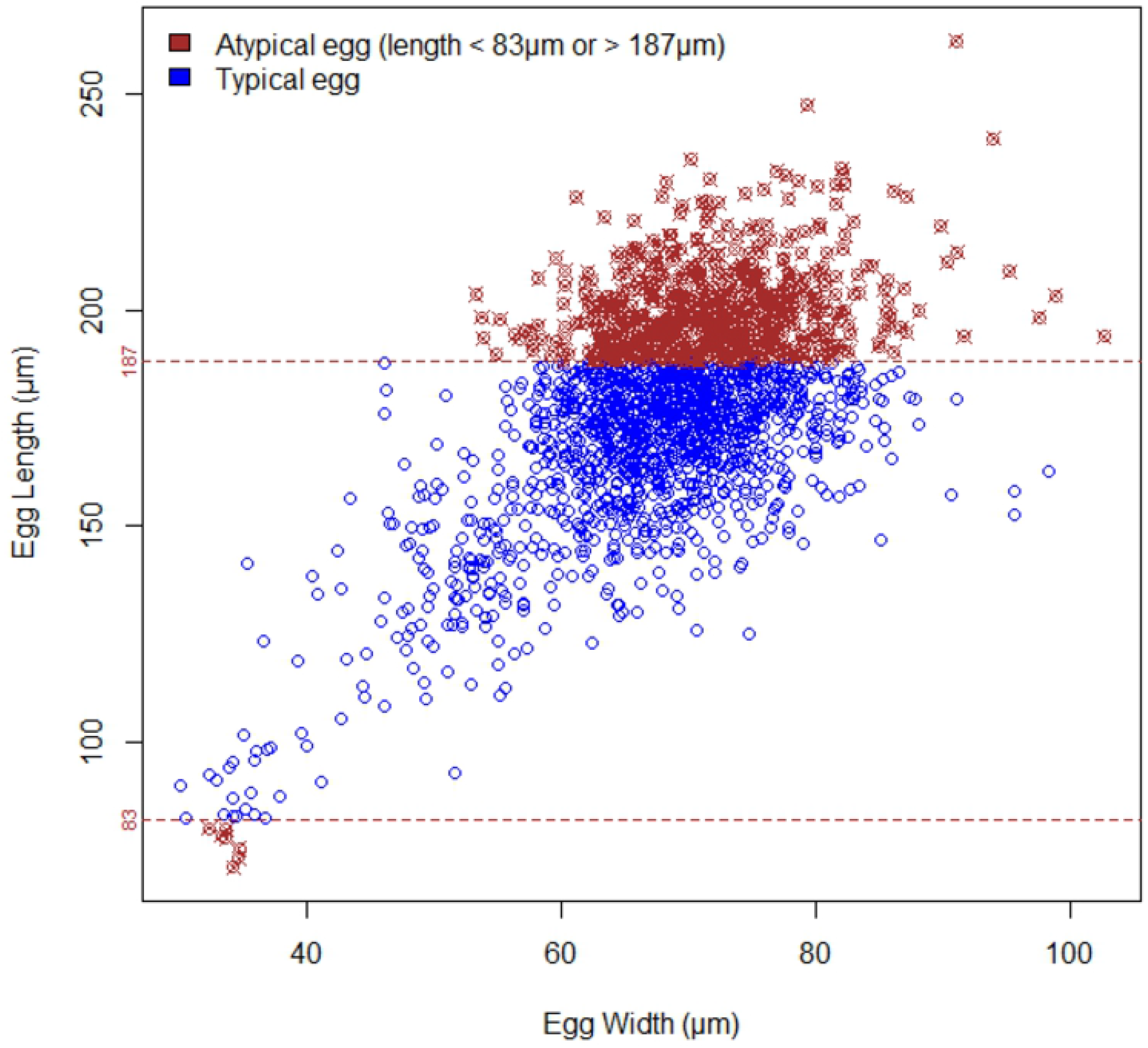
Scatterplot distribution of Schistosoma egg morphotypes recovered from human urine.

### Molecular characterization of *Schistosoma* eggs

Fifty-four (62.1%) of 87 schistosome eggs showed amplification of the schistosome *Dra1* gene (Table 4) (Fig 7). Out of the 54 DNA samples, which were further subjected to PCR amplification of the schistosome ITS-2 rDNA, thirty-three (61.1%) showed varying differences in product sizes (Table 4) (Fig 8). Eight representative samples with different band sizes were selected for sequencing. Nucleotide sequence BLAST search with the NCBI database showed that one egg from an infected person from Abule-Titun was confirmed to be *Schistosoma magrebowiei* with its nucleotide sequence BLAST giving a sequence homology of 100.0% similar to the Zambian strain of *Schistosoma magrebowiei* (accession number UZAI01000474.1). Another schistosome egg from an infected participant from Ibaro-oyan showed similarity to *Schistosoma japonicum* with its nucleotide sequence BLAST giving a sequence homology of >75.40 % similar to the isolate HuSjv2 of the Asian *Schistosoma japonicum* from China (accession number SKCS01001458.1). Both sequences have been submitted to the NIH GenBank database and were assigned accession numbers MZ451359 and MZ451360 respectively (Table 5).

**Table 4:** PCR amplification of schistosome Dra1 and ITS-2 subunit genes.

|  | Number of eggs<br>screened | Number of eggs<br>showing<br>amplification (%) | Number of eggs showing no<br>amplification (%) |
| --- | --- | --- | --- |
| PCR (*DraI*) | 87 | 54 (62.1) | 33 (37.9) |
| ITS-2 | 54 | 33 (61.1) | 21 (38.9) |

**Table 5:** Blast results.

| Source of schistosome egg sample | Location of sample | DNA blast identification | Accession Number | Maximum Identity (homology) |
| --- | --- | --- | --- | --- |
| Urine | Abule-Titun | *S. magrebowiei* | MZ451359 | 100.0 % |
|  | Ibaro-oyan | *S. japonicum* | MZ451360 | >75.40 |

**Fig 7.**
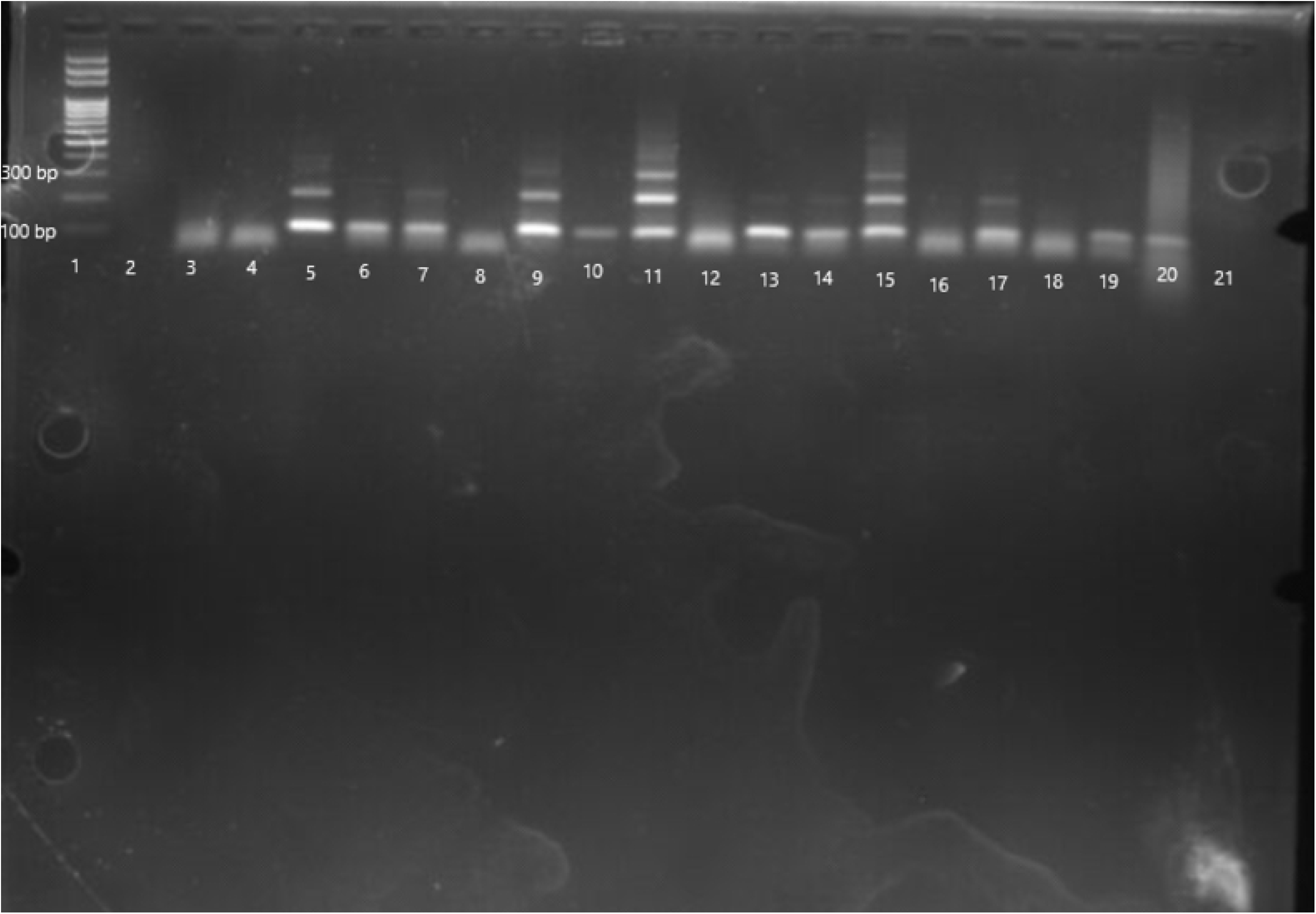
**PCR amplification of schistosome Dra1 as shown by agarose gel stained with ethidium bromide. Lane 1: 1000bp marker, lanes 2-4, 6, 8, 10, 12, 16, 18, 19: no amplification, lanes 5, 7, 9, 11, 13-15, 17: amplification, lane 20: positive control, lane 21: negative control**.

**Fig 8.**
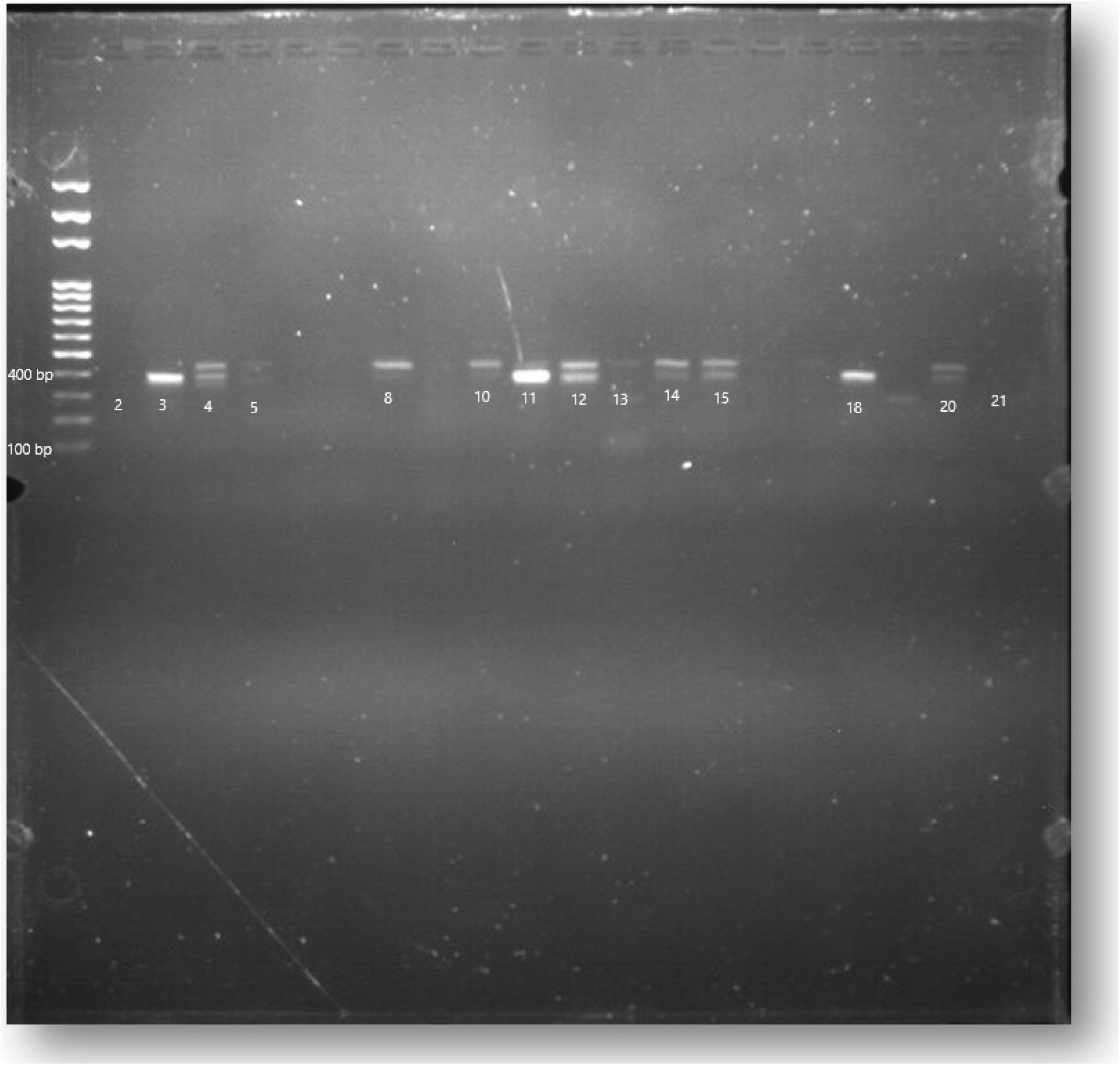
**PCR amplification of schistosome ITS-2 as shown by agarose gel stained with ethidium bromide. Lane 1: 1000bp marker, lanes 2, 6, 7, 9, 16, 17: no amplification, lanes 3-5, 8, 10-15, 18 and 19: amplification, lane 20: positive control, lane 21: negative control**.

## Discussion

An overall prevalence of 57.0% was recorded for urinary schistosomiasis across the study communities, with a range between 47.1% and 63.8%, hence falling within the WHO hyperendemic thresholds [2]. These findings conform with earlier reports, highlighting unabated transmission of schistosomiasis across study area, despite ongoing chemotherapy interventions [11,12]. We demonstrate, for the first time, different morphotypes and morphology of *Schistosoma* eggs in Ogun State and Nigeria. Our findings showed polymorphism of the egg morphotype, with 32.2% of the schistosome eggs comprising the atypical spindle-shaped group. Previous studies have reported the presence of two or three morphotypes of schistosome eggs recovered from both infected humans and animals [21,22,23]. The presence of the different morphotypes in the schistosome eggs recovered from human urine samples in Ogun State suggests hybridization among the *Schistosoma* groups in the area. The schistosome eggs’ lengths were 70.90 – 262.30 μm with a mean length of 176.97 ± 0.53 μm while the widths were 30.10 – 102.60 μm with a mean 68.25 ± 0.20 μm. the mean length in this study correlates with past findings done on the morphometry of schistosome eggs such as [17] (83-187 μm) and [22] (148-223 μm). However, the mean length exceeded that of previously reported findings [16,18,21] and this may be due to the difference in the number of schistosome eggs measured.

PCR amplification of the *Dra1* gene showed the expected tandemly repeated bands in 62.1% as 37.9% of the samples did not. The reduced number of amplicons in this might be due to the low concentration of DNA materials as a result of the phenol-chloroform extraction method used. The sequencing analysis of the amplified ITS-2 showed the occurrence of Asian *Schistosoma* species, *S. japonicum* and the animal schistosome, *S. margreboiwie* in two of the study areas and more confirmatory data is needed to confirm these findings. This is the first report of human infection with *S. magrebowiei* in Nigeria. It is a schistosome species that infect animals such as the Lechwe [24] and a few spurious human cases [25]. It is known to have limited geographical distribution with reports from the Democratic Republic of the Congo, Mali, Zambia, Chad, Botswana, and South of West Africa [26] and host specificity [27] but the immigration and emigration of cattle in the study area due to the activities of nomadic herders may account for the emergence of *S. magrebowiei* in the location surveyed. Our finding shows zoonotic infection of humans with *S. magrebowiei* in the area and suggests possible interspecific interactions with other human schistosome species that may be present in the infected individuals.

Also, we report the first suspected case of the Asian schistosome, *S. japonicum*, in Nigeria. This species is historically known to show a restricted geographical distribution limited to China and other endemic Asian countries [28]. Recent relations between Nigeria and China have spurred the migration of Chinese nationals to Nigeria. This may equally drive the transport of the *S. japonicum* to new areas in Nigeria as human and animal migration have been known to cause the establishment of parasites in new regions in which they did not previously occur [29]. Similarly, cases of *S. haematobium* infection have been found in returning China immigrants from Africa [30]. The transport of *S. japonicum* and *S. haematobium* between Nigeria and China can lead to the establishment of these parasites in allopatric areas if suitable intermediate hosts are present [31].

The findings from our study confirm the coexistence of different schistosome species in the same areas, which infect the same definitive hosts by breaking down physiological barriers and possibly causing mixed infections [32]. High variability in the genetic makeup of *S. haematobium* has been reported in Mali and Nigeria [33] and this may be due to varying levels of interactions between the coexisting species and genotypes in the area [30]. The occurrence or coexistence of different schistosome species in the host may alter virulence and shape the clinical outcomes [34a], presenting novel challenges to existing control strategies and tools by causing high transmission potential and reducing drug efficacy.

## Conclusion

This study was conducted to establish the possibility of hybrid and zoonotic schistosomes in a putative hybrid zone in Ogun state. This study also showed the existence of polymorphism in the morphotypes and morphometrics of schistosome eggs captured. The presence of Asian *Schistosoma* species, *S. japonicum* and animal schistosome, *S. magreboiwei* through DNA sequences call for further monitoring in the transmission of the diseases in the study areas and the state as a whole. Hence, vector snail studies are required to establish the presence of the snail intermediate hosts for zoonotic and hybrid schistosomes, molecular point of care diagnosis (POC) specific to *S. japonicum* and *S. magrebowiei* should be used for screening the communities to establish the burden of Asian and zoonotic schistosomiasis. More studies are required to establish the extent of zoonotic schistosomes in the study areas and the entire state and the survey of other reservoir hosts such as rodents and other small mammals within the endemic communities for cases of hybridization within the *Schistosoma* group should be conducted.

## Data Availability

All relevant data are within the manuscript and its Supporting Information files.

## Acknowledgements

We are grateful to the community leaders and residents of the study sites for their continuous participation. Appreciation goes to the Neglected Tropical Diseases Department of the Ogun State Ministry of Health for providing praziquantel used in treating the positive participants. Our profound gratitude goes to the Nigerian Institute of Medical Research, Yaba Lagos for their support and collaboration in this study.

## Declarations

### Consent for publication

Not applicable

### Availability of data and materials

The datasets used and/or analyzed during the current study are available and attached as supplementary files

### Competing interests

The authors declare that they have no competing interest.

### Funding

Not applicable

## Authors’ contribution

UFE, AFA, AOK and BAA conceptualized the study. BAA, UFE and AFA prepared the protocol while AOP and GVP improved the protocol. BAA, OOO and UCU participated in field surveys and data collection. BAA, OOO, UCU handled laboratory analysis of specimens. BAA, OOO, MHO and OSN performed all statistical analysis. BAA, OOO, UFE and AOP prepared the first draft of the manuscript. All authors contributed to the development of the final manuscript and approved its submission.

## Notes

### Competing Interest Statement

The authors have declared no competing interest.

### Funding Statement

The authors received no specific funding for this work.

### Author Declarations

Ethical clearance for this study (SHA/RES/VOL.4/154) was obtained from Ogun State Hospital, Ijaiye health ethics review board.

